# A mixed methods analysis of participation in social contact surveys

**DOI:** 10.1101/2022.01.28.22270006

**Authors:** Emily Nixon, Taru Silvonen, Antoine Barreaux, Rachel Kwiatkowska, Adam Trickey, Amy Thomas, Becky Ali, Georgia Treneman-Evans, Hannah Christensen, Ellen Brooks-Pollock, Sarah Denford

**Affiliations:** School of Biological Sciences, University of Bristol, Bristol, UK; Bristol Veterinary School, University of Bristol, Bristol, UK; CIRAD, UMR INTERTRYP, F-34398 Montpellier, France; INTERTRYP, Univ Montpellier, CIRAD, IRD, Montpellier, France; School of Population Health Sciences, University of Bristol, Bristol, UK; NIHR Health Protection Research Unit in Behavioural Science and Evaluation, University of Bristol, Bristol, UK

**Keywords:** social contact surveys, epidemic modelling, infectious disease, mixed methods, focus groups, research engagement

## Abstract

**Background:** Social contact survey data forms a core component of modern epidemic models: however, there has been little assessment of the potential biases in such data.

**Methods:** We conducted focus groups with university students who had (n=13) and had not (n=14) completed a social contact survey during the COVID-19 pandemic. Qualitative findings were explored quantitatively by analysing participation data.

**Results:** The opportunity to contribute to COVID-19 research, to be heard and feel useful were frequently reported motivators for participating in the contact survey. Reductions in survey engagement following lifting of COVID-19 restrictions may have occurred because the research was perceived to be less critical and/ or because the participants were busier and had more contacts. Having a high number of contacts to report, uncertainty around how to report each contact, and concerns around confidentiality were identified as factors leading to inaccurate reporting. Focus groups participants thought that financial incentives or provision of study results would encourage participation.

**Conclusions:** Incentives could improve engagement with social contact surveys. Qualitative research can inform the format, timing, and wording of surveys to optimise completion and accuracy.

**Graphical abstract:** 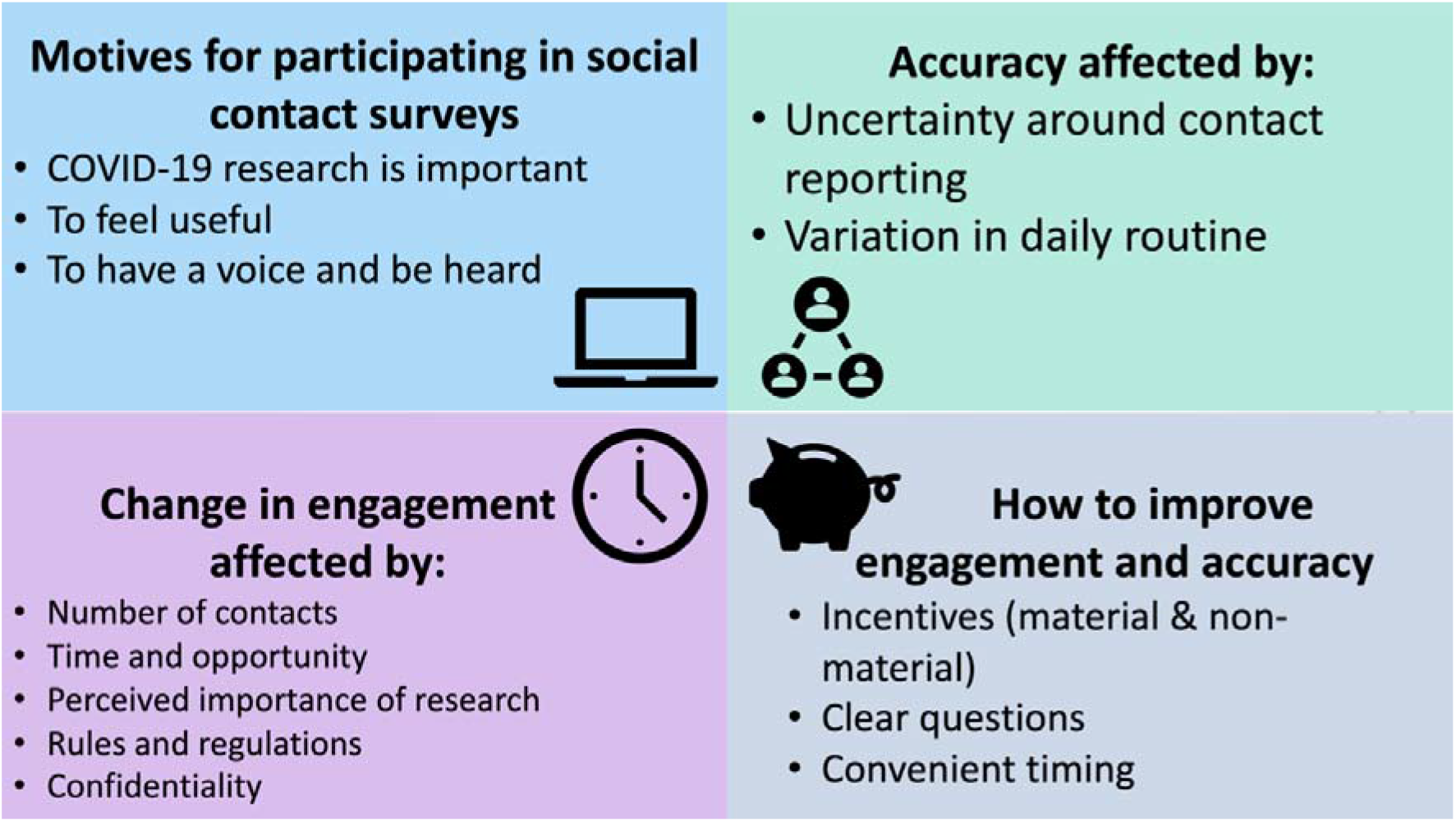

## Introduction

Social contact patterns, or who-meets-whom, are central for understanding and quantifying the transmission of airborne and close contact infectious diseases such as COVID-19, influenza and measles. Social contact patterns determine how rapidly a disease propagates through a population, which individuals are most affected and the likely impact of interventions (1).

Collecting empirical data on social contacts is challenging because of the transitory and nebulous nature of a social contact. Whereas data on sexual contacts have been collected and used since the 1990s, measuring social contacts was initially considered too unreliable. The first study to demonstrate the useability of social contact data defined a social contact as any two-way conversation (2); this definition of a social contact has been refined in various ways to require at least 3 words, or to not involve a raised voice or to be within 2 metres of the other person (3). Studies have shown that age is a key determinant of numbers of social contacts with younger people reporting higher numbers of contacts and strong age assortative patterns (individuals tend to contact others of a similar age) (4,5).

Most social contact surveys ask respondents to list all social interactions on a single day, either retrospectively or prospectively (6). The average number of individual contacts reported on a single day is approximately 10, however there is large variability between individuals, with evidence suggesting this is closer to 20 on a week day among university populations(7–9). The Social Contact Survey demonstrated that when respondents could report groups of contacts, the number of contacts was considerably higher (10), suggesting that there may be a disincentive for reporting high numbers of contacts individually.

There are inherent biases in participation in studies such as social contact surveys. Women are more likely to participate than men (11) and lower rates of participation have been associated with lower educational status, smoking, young age, lower vaccine rates and living with children (12). For studies that require regular or continued participation, women are less likely to be high participation users (13) and online recruitment may lead to lower long-term participation rates (14). On top of biases in participation, some types of contact are less likely to be reported, for instance short duration contacts (15) and weekend contacts (16).

In this paper, we present a mixed methods analysis of our experience implementing a social contact survey in a university setting during the COVID-19 pandemic (CONQUEST: Contact patterns and behaviour in University of Bristol (UoB) staff and students during the COVID-19 pandemic). This is an ongoing, weekly, longitudinal survey which has been established since 23rd June 2020. Participants are asked to retrospectively report yesterday’s contacts, split by individual and group contexts. Quantitative results from the survey have previously been published (17,18). Here we adopt a mixed methods design to a) explore student participants’ experiences of taking part in CONQUEST, b) aid understanding of the quantitative data collected through the survey and c) identify factors that might influence the validity of survey results. As qualitative research in general focuses more on descriptive questions, it can provide broader contextual understanding to complement quantitative data (19). Whilst quantitative data were able to determine crucial information regarding student contact during the COVID-19 pandemic, it is unclear exactly how students were interpreting and answering survey questions. Qualitative data can provide detailed insight into participants’ interactions with the survey: their experiences of, motives for, and potential problems with survey completion. These insights provide important context for interpreting outputs from contact surveys and evaluating the validity of survey responses and will help with the design of future contact surveys which inform epidemic models.

## Materials and methods

### Qualitative methods

#### Design

Qualitative online focus groups were conducted to provide insight into how and why people were taking part in COVID-19 contact surveys, and what could be done to improve participation and retention. Face-to-face data collection was not possible due to social distancing restrictions at the time. Focus groups were chosen as a method of generating data because, within this group dynamic, participants are encouraged to explore issues, identify common problems, and suggest potential solutions through sharing and comparing experiences. As Morgan (1996) states (20), groups can provide comparisons that lead people to talk about a wider range of experiences and opinions, more so than those that occur in individual interviews. Interactions in groups can provide “more accurate accounts of what people actually do” (ibid., p.232). Also, the researcher-researched relationship changes within focus groups and can “shift the balance of power in favour of the participants” (21) leading to more natural discussions and sharing of rich data within a setting less rigid than interviews. The focus group method also enabled the researchers to gather a large amount of data in a relatively short period of time.

#### Recruitment

All participants were required to be students at the University of Bristol. We did not recruit University of Bristol staff as we were particularly interested in behaviours in population groups with a high number of contacts. We also had had less participation from students in CONQUEST and so we wanted to find out more about to engage this underrepresented group. To recruit individuals for the focus groups who had previously taken part in CONQUEST, we sent an email to a mailing list of all participants who had consented to be contacted about further research. This mailing list included those who regularly completed CONQUEST and those who were no longer completing or only rarely completing CONQUEST but had not opted out of the mailing list. To recruit those who had never completed CONQUEST, a study advertisement was circulated via University of Bristol social media accounts, university newsletters and on the university portal. In all advertisements for the focus groups, it was mentioned that participants would be given a £20 shopping voucher for one hour of their time.

Interested participants were directed to an online information sheet about the study. Those willing to take part were asked to consent to participate via an online “tick box.” This enabled access to a pre-screening questionnaire (hosted by Microsoft Forms) in which participants were asked to provide basic demographic and contact details, as well as to indicate their availability and whether they had (n=38) or had not (n=36) completed CONQUEST. Participants were divided into four groups, based on whether they had completed (2 groups) or not completed (2 groups) CONQUEST and when possible, whether they were a postgraduate or undergraduate. Participants (n=42) were selected based on their availability and on key demographic characteristics (faculty, year of study, international status, sex, ethnicity, age) with the aim of having the most diversity possible in each focus group. Only 27 of the selected participants attended the focus group sessions. There were four focus groups and group sizes varied from 5 to 8 participants.

#### Data collection

Focus groups took place on a video call in September 2021 and lasted up to an hour. These were moderated by a qualitative researcher from the University of Bristol who was responsible for asking questions and leading the discussion. An assistant moderator was also present to take notes, ensure equal involvement among participants, and monitor the online chat function. The assistant moderator was available to provide support to anyone who became distressed by the discussions should it have been required.

Each focus group followed a semi-structured topic guide (see supplementary materials) with questions designed to explore why people were or were not regularly completing the CONQUEST survey, why people may or may not take part in various research during the pandemic, and what could be done to improve completion and retention. Toward the end of the focus group, students were asked to comment on specific questions about reporting of contacts, including their interpretations of the questions, and how they (and others) may have answered those questions.

With permission, focus groups were recorded and transcribed verbatim. All participants received a £20 Love2Shop voucher as reimbursement for their time.

#### Analysis

Data were analysed using a thematic approach (22). Following the six stages of thematic analysis, data were read and re-read by two authors (TS and SD). Codes were then assigned to dataset independently by the same two authors, examined and collated into potential themes. Themes were then reviewed and checked against the data to ensure they are a convincing interpretation of the focus group discussions. The team met regularly throughout this process to discuss the themes and ongoing analysis.

#### Ethics

Ethical approval was obtained from the Faculty of Health Sciences Research Ethics Committee at the University of Bristol (application reference number 116774). All participants of the focus groups and participants of CONQUEST were given information about the study before they gave informed consent.

#### Quantitative methods

After completion of the thematic analysis of the focus groups, we conducted complementary quantitative analyses using data from the CONQUEST survey itself. The aim of the quantitative analysis was to provide context to the qualitative analysis and highlight links between themes discussed and the subsequent impact on survey responses. All analyses were completed using R version 4.0.4, other than the analysis on one-time participants versus repeat participants which were completed using STATA version 16.1. To compare contacts of repeat and one-time participants, a negative binomial regression model with the number of contacts as the outcome was used on a dataset containing the survey responses (2020/06/23–2022/01/24), adjusting for whether the answers were from a repeat respondent or one-time respondent, age (transformed using cubic splines), sex, post-graduate status, and month of response. We used publicly available data on University of Bristol student demographics (23), anonymised data from the form completed by focus group participants and anonymised data from the CONQUEST survey.

## Results

### Demographic characteristics of the focus group participants

There were 27 participants across the four focus groups, 13 of these had never completed CONQUEST and 14 had completed it more than twice. All faculties from the university were represented by our participants, including the Faculty of Arts (n = 5), Faculty of Engineering (n=3), Faculty of Health Sciences (n=5), Faculty of Life Sciences (n=3), Faculty of Science (n= 5), and Faculty of Social Sciences and Law (n=6). We had a lower proportion of undergraduate students (0.59) than seen in the university population or in the CONQUEST survey (Figure 1, 0.74 and 0.66 respectively) and, therefore, a higher proportion of postgraduates (0.41). Of the 16 undergraduate students, 5 of these were first year students, 4 were in their second year, 4 in third year and 3 in fourth year. Of the 11 postgraduate students, 7 were in their first year, 3 in their second year and 1 in their fourth year. Only 1 international student took part in the focus groups. Almost three times as many participants in the focus groups were female (n=20) than male (n=7), which is a similar proportion of female students (0.7) to those taking part in the CONQUEST survey (0.68), but higher than the total proportion of females in the total university student population (0.55) (Figure 1). A higher proportion of students with white ethnicity (0.85) took part in the focus groups, compared to the 0.74 taking part in CONQUEST and the 0.63 in the total university student population (Figure 1). Our youngest participant was 19 years old; the oldest was 55. The median age of participants was 22 years.

**Figure 1.**
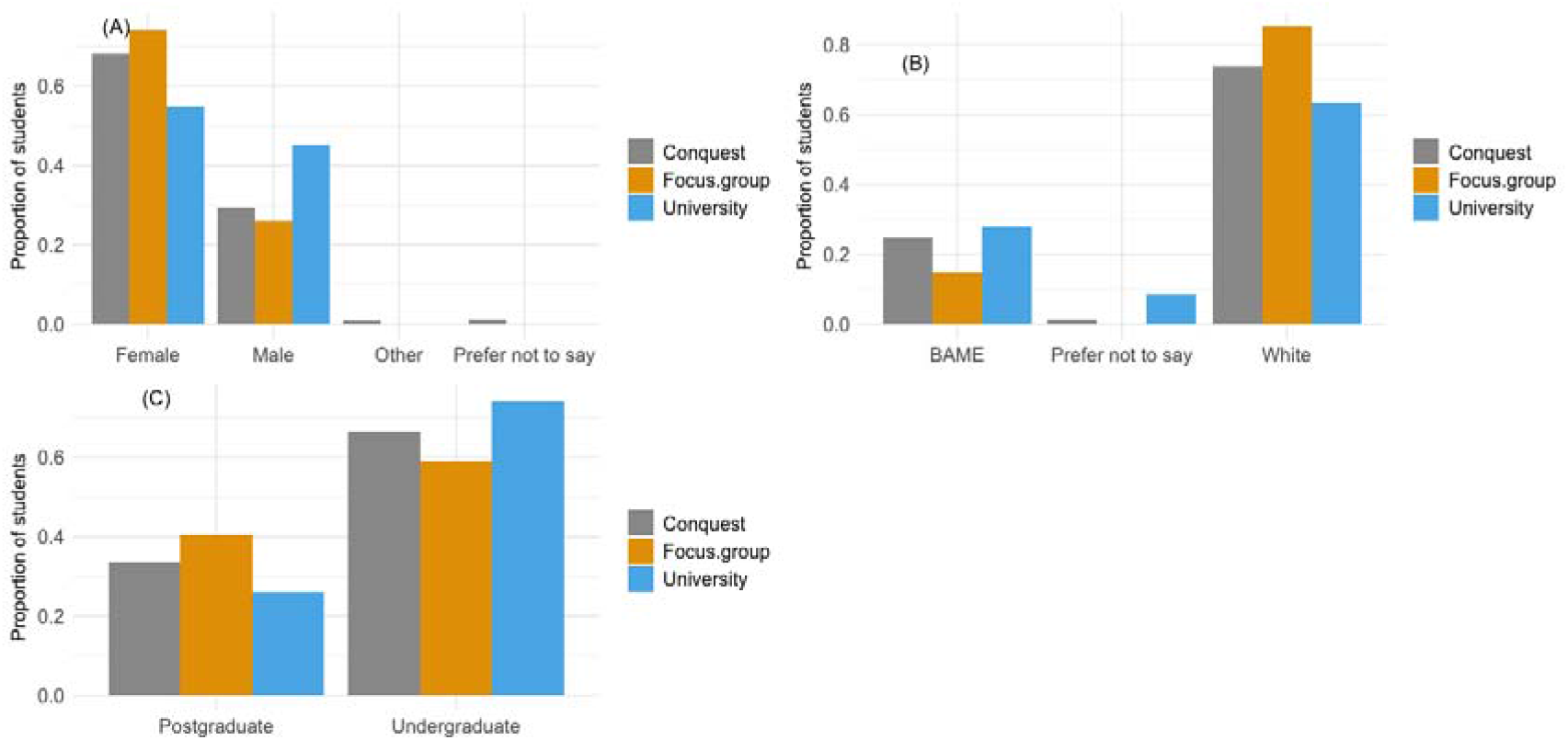
Demographic of students, taking part in the focus group or completing CONQUEST until the 6^th^ December 2021 compared to the overall demographics of UoB students. (A) sex, (B) ethnicity, (C) graduate status.

### Motives for taking part in social contact surveys

Students provided three main reasons underpinning their decision to take part in social contact surveys: 1) COVID research is important 2) to feel useful 3) to be heard.

### Covid research is important

Participants described being motivated to take part in research that they considered to be important. Due to the huge impact of the pandemic on the lives of the population, COVID-19 research was considered to be of the highest importance. Students were willing to increase their efforts and spend longer completing research if it was considered important enough:

*“It [CONQUEST survey] is long, but if it is presented as something that is important – it’s something that’s representative of a global happening that is unprecedented, and someone’s been given an opportunity to participate in, in assembling data about it – so it’s manageable because it’s important” (P2, G4)*.

Students thought it important to provide accurate data that reflected real world events:

*“It’s more important to know what people are actually doing, so acting based on what the laws are and basing policy off that isn’t necessarily very helpful because it might be completely inaccurate to what people are actually doing (P4, G2)*

### To feel useful

Students described feeling “less useful” during the pandemic, and participating in research was viewed as an opportunity to contribute:

*“For me, it was about feeling useful at a time where we were all working from home and I certainly didn’t feel as useful as I had previously” (P6, G1, took part in CONQUEST)*

The research provided an opportunity for students to help and potentially contribute to research that could inform the national response to the pandemic:

*“I just wanted to try and contribute to research to inform public policy” (P6, G2)*

Students who may otherwise have been reluctant to take part in research were willing to do so for the greater good:

*“It also makes you feel like you are helping…I don’t really like giving my data away, but I felt like if it was used for some kind of social good, you kind of feel a bit more comfortable with it” (P1, G3)*.

Rising cases of COVID-19 in the student community reinforced the need to help to end the lockdowns:

*“The time I first got [the invitation to complete CONQUEST], it was one of the peaks, when everyone was catching COVID… I suppose, in my mind, it was like, ‘every little helps. If you can do this and then they figure something out from it in some way which is going to make lockdown end quicker then that would be great*.*’, so I was doing it to get through things quicker” (P3, G2)*

### To have a voice and be heard

Participants noted the need for research to include and spotlight student populations, providing a chance to be heard:

*“You definitely needed this demographic to talk” (P8, G1)*.

It was acknowledged that student communities are unique in their interactions, and research conducted with general populations may not always be relevant to them:

*“Students don’t behave necessarily in the same way as maybe a household of parents and children do, and I feel like I wanted to do it, I guess, to contribute to making a more accurate picture of how students might engage with other people in a pandemic” (P5, G2)*

Students felt they were often villainised, and wanted an opportunity to demonstrate that this was unjust:

*“There are a lot of students that are seeing hundreds of people and are probably feeling guilty about it, and they see in the media… that campaign that was, ‘Don’t kill your nan’ – that kind of thing. So, maybe if you appeal directly to that and show that young people aren’t causing a spike in the pandemic, and they were trying to collate data on young peoples’ activities to demonstrate that they’re not ‘the bad guys” (P4, G4)*.

### The role of changing levels of restrictions on survey completion

The level of student participation in CONQUEST has varied greatly across the study period (Figure 2). Recruitment campaigns increased participation; however, the recruitment campaign in 2020 enlisted around five times more participants than when the same recruitment campaign was run in 2021. A decrease in student participation in CONQUEST was seen after all three lockdown periods across 2020 and 2021.

**Figure 2.**
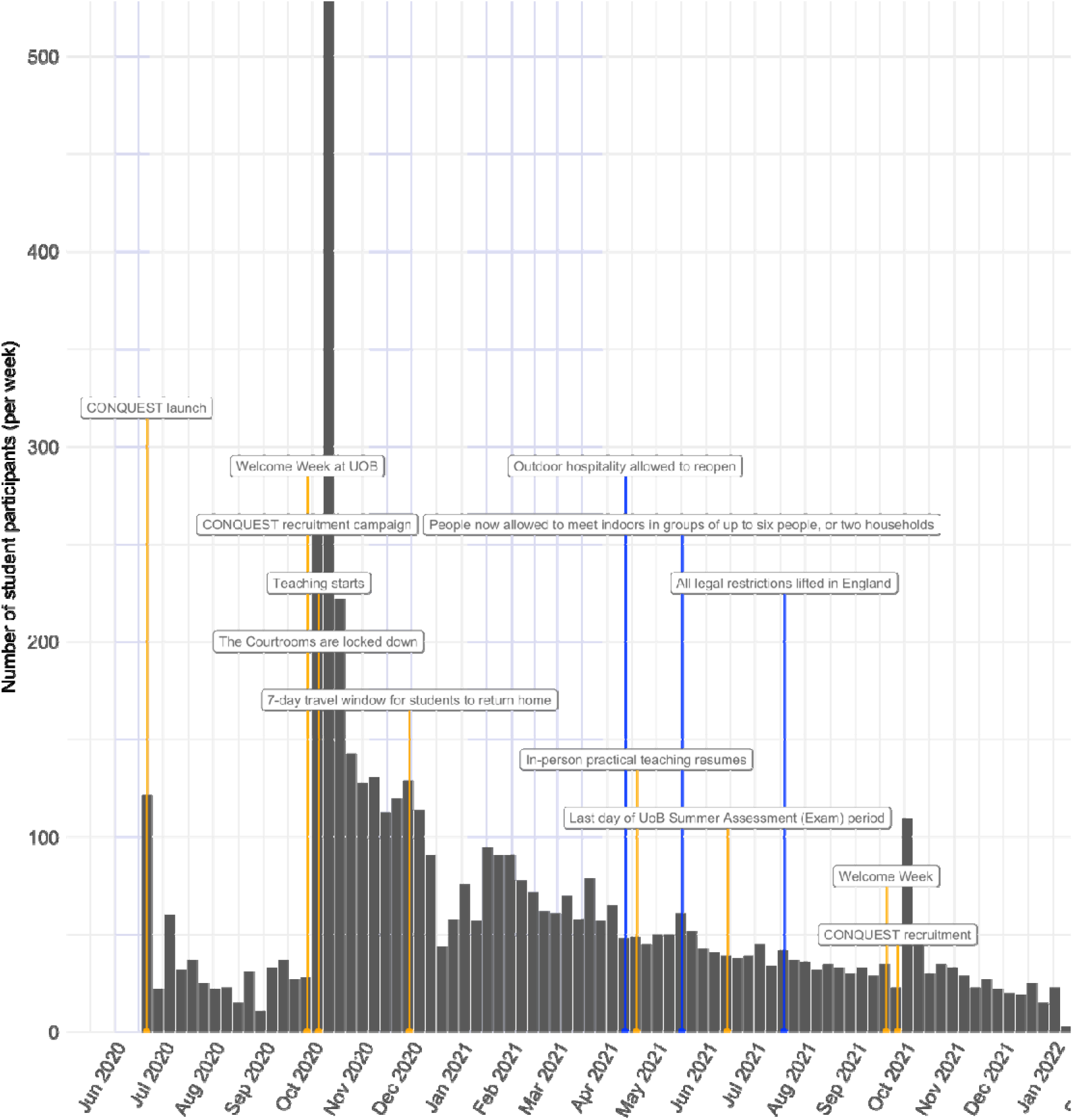
The number of students completing CONQUEST each week. University milestones specified as orange labels and easing of national restrictions as blue. The areas shaded in grey indicate the months where a national lockdown was in place.

CONQUEST participants who completed the survey more than once had more reported contacts across their survey responses (N=4620, mean contacts=13.4) than non-repeat participants (N=496, mean contacts=7.1), a result that held in the regression model upon adjustment for age, sex, post-graduate status, and month; rate ratio 1.44 (95% confidence interval: 1.28-1.61; p<0.001). They were also more likely to be older; mean=25.1 (24.4-25.8) vs 22.6 (22.1-23.2) years (p<0.001), and post-grads (48% vs 32%, p<0.001), but were similarly likely to be male (30% vs 29%, p=0.544).

Participants of the focus groups discussed several factors that affected their engagement with CONQUEST, many of which appeared to have been directly impacted by the introduction and removal of social distancing including: 1) number of contacts; 2) time and opportunity; 3) interest and importance of research; 4) rules and regulations.

### Number of contacts

With imposed restrictions of movement and contact, participants often described having a low number of contacts. This made completion of the survey quick and easy, giving participants little cause to stop:

*I’ve probably been at home, I haven’t seen any groups – I’ve only spoken to my boyfriend. It literally doesn’t take any time at all, so why wouldn’t I carry on doing it? (P7, G1)*.

However, as social distancing measures were eased, students would often have more contact with others. This increased participant burden, which could deter participants from completing the survey:

*As things are opening up a bit more and coming into contact with more people, and I think, ‘Oh, God, I’m gonna have to put 12 people on my list of contacts*.*’ Pure laziness, but it is a slight deterrent” (P2, G1)*.

Removal of restrictions also meant participants were coming into contact with new individuals, about whom they may not know key details:

*“I stopped after a while because I think it was when the lockdowns really weren’t that heavy and I was seeing quite a few people each day so it took a really long time to write out every single person’s name and then all their university information which I sometimes didn’t know (P1, G2). It was thought that contact with a high number of individuals could result in participants not filling out the survey, or not including all contacts: “Personally, I think I would just lie ‘cause, once I’ve started a survey, I probably wouldn’t want to stop doing it. I’d probably find it easier to just maybe lie about how many people I’ve seen” (P6, G4)*.

### Time and opportunity

Through the entire study period to date (23rd June 2020 – 5th January 2022), most (56%) of student CONQUEST questionnaires were completed in the morning (07:00-11:59). Lunchtime (12:00-13:59) also was a popular time to complete the questionnaire; considering it was at least half the number of hours as the afternoon (14:00-17:59), a similar percentage of questionnaires were completed (12% and 15% respectively). Students were still completing the questionnaire in the evening (18:00-23:00, 8%) and at night (23:00-06:59, 9%). The pattern of these results did not change much over time (Figure 3), with the morning still being the preferred time to complete the survey (monthly range 40-71%).

**Figure 3.**
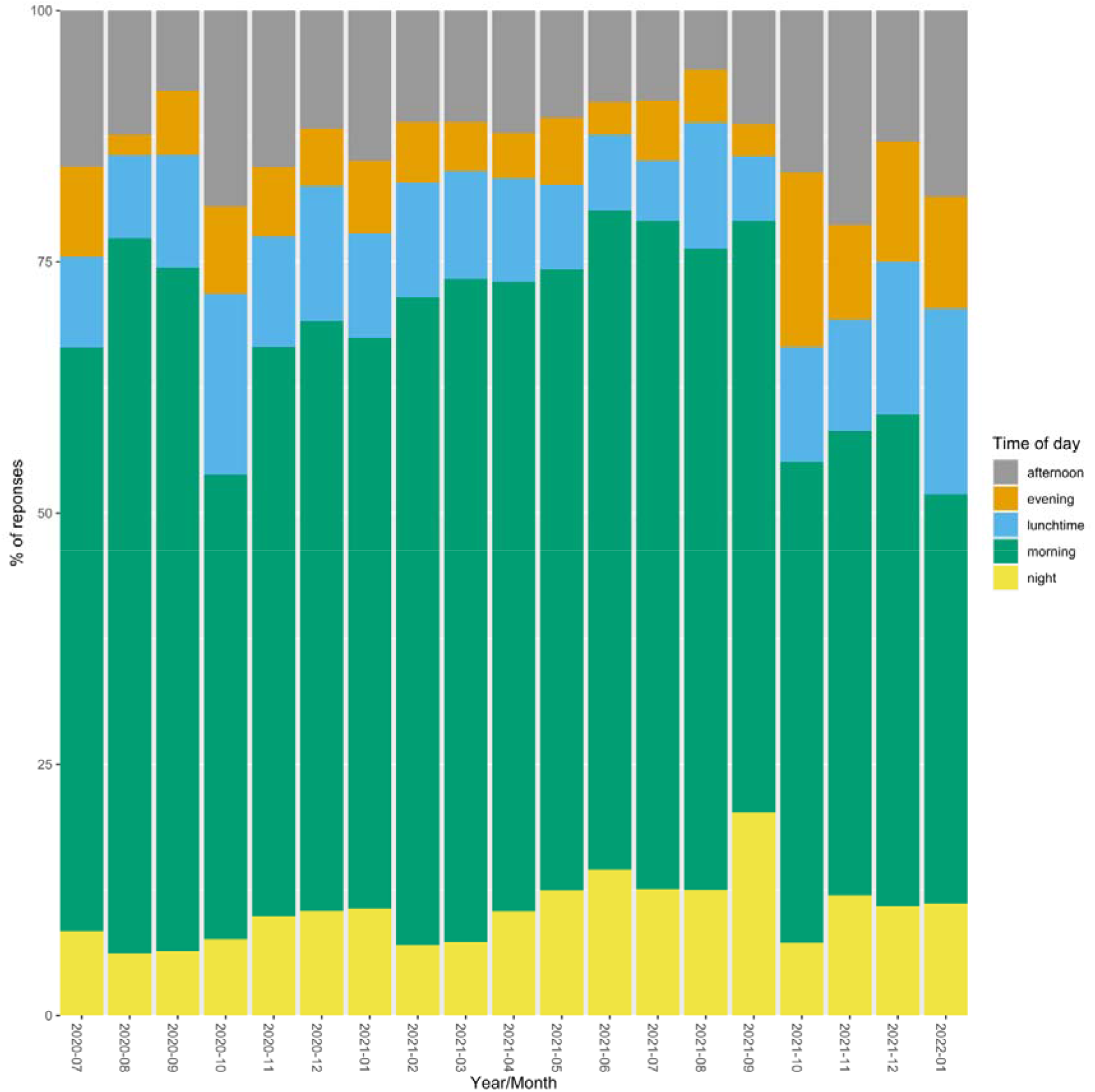
The proportion of students completing the CONQUEST survey stratified by the time of completion: afternoon, evening, lunchtime, morning or night between 23rd June to 5th January.

During lockdown the amount of time the UK public spent on electronic devices increased considerably as people were required to stay in their places of residence more. Online surveys were therefore much easier to complete:

*“The way that we’re working at the moment with lots of Zoom meetings lends itself well to just quickly doing the survey but, as that changes – perhaps as we are on campus more – I might be inclined to do it less. I’m not sure” (P6, G1)*.

Zoom meetings in particular provided an ideal time to complete a survey:

*“If it turns up when I’m in boring Zoom meetings, then I always fill it out because I’m on my laptop and I’ve nothing better to do! I have Zoom meetings at 10:00am most days! The 10:00am slot is a good one!” (P4, G1)*.

Focus group participants reported that online surveys were easier to complete whilst they were spending most of their working time online. However, as lockdown restrictions were lifted, circumstances changed, and opportunities for completing surveys reduced. At the same time, the number of possible contacts increased, thus increasing the time needed to complete the survey:

*“One thing I noticed is that when lockdown restrictions were eased or not as strict and I was seeing more people, I was then also busier and then didn’t fill it out because it was a longer job, but I was also out doing more things so when I was seeing more people I was less likely to fill it out anyway” (P1, G2)*.

### Impact of lockdown on sustained interest / importance

An easing of restrictions appeared to reduce the importance of COVID-19 and associated research, with students keen to return to life as normal:

*“I think maybe just with everything easing, you start thinking about COVID a little bit less, so maybe feel like it’s less important to do now that everything’s getting more relaxed” (P3, G1)*.

### Rules and regulations

Students described how concerns about the possible consequences of breaches of regulations caused them some unease and could prevent them admitting to contacts that were not permitted at the time, particularly if breaches were substantial:

*“If students were to fill it in and lots of students haven’t been keeping so much to the COVID guidelines. There might be a fear of students filling it in*… *if you’re a Fresher, you’re in contact with hundreds of people, so I don’t know if there’s a guarantee of not reporting them to the police, or you’re allowed to say whatever you want kind of thing, because I think that would probably put a lot of students off because they’re not gonna say, ‘To be honest, I’ve been in contact with 300 people since the beginning of the year’ because they know it’s illegal” (P4, G4)*.

*“I was very hesitant to say if I’d been on a protest because police were seeing it as illegal but more illegal than they were seeing breaking COVID regs, so I was very hesitant” (P4, G2)*.

Even though students were aware that the survey was anonymised, an element of doubt remained:

*“I still fill it out anyway, and think, ‘It’s okay – it’s anonymised anyway*.*’ It’s a little bit of a nagging thing there” (P3, G1)*.

Whilst those with medical backgrounds were aware of anonymity procedures, it was thought that this may not be clear to others:

*“I guess I knew [that data would be anonymous] from the research papers which I’d read as part of my degree, but I think for other people, maybe if you didn’t do a life science or a physical science then you wouldn’t have known that so I guess it’d be really easy to assume that you’d be seen, so I think making that explicit would be really nice” (P2, G2)*.

It was noted that the fact that the survey was perceived to be run by the university (rather than by a research group at the university) may have exacerbated these concerns:

*“This is one where actually the relationship with the University maybe isn’t the best, because particularly students living in halls, I think they would be a little bit concerned about declaring that [they have and what implication is… although they probably know deep down it wouldn’t be passed on and although all the information about data sharing is published, I feel a bit uncertain about that” (P2, G3)*.

Those with legitimate reasons for having a high number of contacts (e.g., through work) were concerned that this may be misinterpreted by their schools or departments, or that they would be criticised for attending classes following increased contact:

*“If my school was able to look at my contacts, they might think, ‘Oh, my God, she’s meeting so many people at the weekends,’ whereas, actually, I’m just going to work. But, if they turned around and said, ‘Well, hang on, you’re going to work on Sunday, and meeting 50 people, and then you’re coming to campus on Tuesday, how do we know you’re not giving us…?’ I’d feel really bad, and I wouldn’t want to have to get into the debate of, ‘Oh, the school don’t like me working ‘cause I could be spreading Coronavirus to the students and the staff’” (P2, G1)*.

Other students reported that the lack of opportunity to justify particular behaviours, or capture any safety precautions that were in place meant that their answers could be misconstrued:

*“I think I’ve been very safe, but that’s not always to the letter just because like the broad rules have to be applicable to everybody, but in specific situations, I think you can be safe without doing exactly as you’re told. If there was a box where when I admit to it I can type out all my justifications, then I would be much more likely to admit to it rather than if it just looked like I was just not caring at all, then I might resent being forced to admit that” (P1, G1)*.

### Factors influencing reporting of contact patterns

Participant discussions revealed a number of ways in interpreting and reporting of contacts may influence data. This included (1) uncertainty around contact reporting and (2) impact of the day of the week

### Uncertainty around contact reporting

A number of students who had been regularly completing CONQUEST requested greater clarity regarding how contacts should be reported. In particular, many requested clarification regarding differences between group and individual contacts:

*“If I’m meeting with flat-mates but if I was living with two other people and we’re in a friendship group of six of us, so the group that we met up with was a group of six but I was living with two of them, so I don’t know if I’d put them under flat-mates because I met them individually or would I put them as part of a group because I met them as a group? Would I put them as both, but then would you count them twice? That I found was more confusing” (P4, G2)*.

Some students were unclear about when a contact became a contact and how to deal with contacts that fall into different categories:

*“When it says, ‘contact you had with other people yesterday including those that you live with’, if you live with someone but, say, don’t see them, would you not mark them down because you didn’t have contact? Is it just contact including those you live with or is it contact ‘comma’ including those that you live with? So if you hadn’t seen them, would you put them down?” (P4, G2)*.

*Also, where to put people who fall into different categories so, for example, I live with my housemate but, every other week, we work with an overlapping shift pattern where we’re in contact with the people we work with, and they’re also a student, but they’re also my housemate. I’m like, ‘Do I have to put them in every single section, depending on what kind of contact I’ve had with them?’ Typically, I’m lazy, and I just put them down as my housemate, and forget that fact that I work with them! because I don’t know if I’m just repeating it or not” (P2, G1)*.

Participants had questions about when they were supposed to be completing the survey:

*“I wasn’t sure whether I was meant to be answering for the day I received the email or the day I read the email so, if it had been like a few days, I thought, ‘Oh, it’ll be out of date, and out of synch, and I’ve missed the time point,’ and I don’t remember that long ago, so I skipped a few weeks because of that” (P1, G1)*.

Among those who had not completed the survey, many were concerned that they did not have the knowledge to answer the questions honestly:

*“I don’t think I’d be able to fill it out. I just don’t remember or don’t completely [know] so yesterday I went to the library, I went to the gym, I don’t completely know how I would fit those in to which section, so yeah I don’t know” (P5, G3)*.

Students had multiple ways of dealing with uncertainty; including stopping the survey (11% of survey responses were only partially completed), or including contacts twice (not always consistently):

*“I think just any questions that are obscure or don’t make sense*…*you can see a badly written question it really puts you off answering it” (P3, G3)*

*“[if I didn’t know where to put the contact] I think I’d probably end up putting them in both” (P1, G3)*

*“I certainly find myself varying week by week whether I put [contacts] in that one at the bottom, or on the next page in the group section. Definitely varies” (P6, G1)*.

Some participants also described strategic behaviour in terms of when they chose to complete the survey. In some cases, this related to their aims to provide more specific information:

*“I’ve tried to show the diversity in my week by postponing it and timing it differently at different times. In general, I do it on a Tuesday, but sometimes I’ve delayed it either because I don’t have time on that particular Tuesday, or because I’m like, ‘Oh, this week is a very different week – I’m gonna do it on a different day’” (P6, G1)*.

This strategic reporting also reflected their concerns about whether the information they were reporting was accurate or whether they were reporting it correctly:

*“I think there was once when I started and then realised that a group I’d met up with – well, it wasn’t a group, it was something niche, I didn’t know quite where to put it in the thing so I think I essentially went, ‘Cool, I’ll do the thing tomorrow, then*.*’, and so I did it tomorrow so I didn’t have to deal with the fact that I didn’t know where to put this thing” (p4, G2)*.

Some also speculated how to avoid reporting several contacts and whether this would defer them from completing the survey in general:

*“*…*if I was seeing loads of people in a week and I knew that was gonna make my survey longer, probably I would either lie and skip people out, or maybe not bother once I realised that every single contact was being recorded, I think it might put me off, but obviously that is the whole survey” (P5, G4)*.

### Impact of the day of the week

The most popular day for students to respond to the survey was Monday (20%), all other weekdays each had 15% of the responses. A lower proportion of responses took place on Saturday and Sunday (8% and 9% respectively).

A number of participants discussed how variable days could be with regard to contacts made:

*“So I was just checking my emails and it came on every Monday for the past four weeks. Doesn’t seem very randomised to me because I have a certain pattern that I do most weeks on a Monday. A different one on a Thursday and, if you asked me on either of those days, my responses would be very different” (P8, G1)*.

Some students would select to complete the survey on their busier days, as these were considered more interesting and more helpful:

*“Sometimes I would wait until I’d had a more interesting day to fill it out if I’d had a stream of boring weeks, but I think the fact that it comes on the same day each week is perhaps not best suited to reflect the diversity of what we do in our week” (P6, G1)*.

*“Cause I worked virtually all the lockdown so, on my days off, it was really easy to fill out because I only saw people in my house when I was in work. I’m a support worker, so I was in work – I had a lot of contact with people. Some days it was easy, some days it wasn’t, but I was quite committed just to continue doing it because I’d signed up for it, and I thought it would be more helpful on the days that it is more difficult ‘cause it’s more information in that” (P5, G1)*.

However, some students felt that having a high number of contacts on a particular day may result in a situation in which honesty and integrity are compromised:

*“I think even with this level of detail and even like what faculty people are from, like questions, brackets like that makes me question what the data, like why is the faculty important and in terms of question fatigue, I think after doing that five times I would be getting quite tired and a normal day I would be meeting more than five people for sure” (P3, G3)*.

*“Most students would probably just lie about how many students they met” (P2, G4)*.

*“I would start filling in a flatmate and then think, ‘Oh, I have to do this for, like, three more other people*.*’, and then I would stop” (P1, G2)*.

### Potential to increased participation through greater student benefit

It was generally thought that provision of incentives would increase participation and sustained engagement in the study, with the incentive being reflective of the level of commitment:

*“I think I mean similar to this call if there is funding to pay people to take part or to receive vouchers, then that’s going to get people” (P3, G3). “Yeah, I don’t think it needs to be much especially if it’s just a small survey, but just any kind of incentive is just something*.*” (P4, G3)*.

*“I think to do a more longer term study where I was committed and giving repeated data, I think I’d be more likely to do it with some kind of incentive” (P3, G3)*

Incentives could be vouchers, cash, or points schemes, although prize draws may be less appealing:

*“I think I’ve seen quite a few surveys or things like that recently and I think it’s definitely an incentive but what I often think is my chance of winning is actually so slim, like what’s the point. So I don’t know if that might go through quite a lot of people’s minds” (P5, G3)*.

For multiple or repeated surveys, participants recommended an accumulative points system:

*“I do YouGov surveys as well and they give you points for each survey that you complete. once you get a certain amount you can redeem a voucher or a bank transfer” (P1, G3)*.

In the absence of cash or financial incentives, personalised feedback would be acceptable:

*“I don’t know about other people, but I love a good statistic! I love to know what everyone else has been up. So, like, ‘The average number of people in your age group come into contact with is this…’ during that day, or just a bit of an insight into some of the data that’s being collected. I don’t know. I found that really interesting – to see if I’m meeting up with 10 times more people than everyone else” (P2, G1)*.

All the main themes identified are outline in Figure 4.

**Figure 4.**
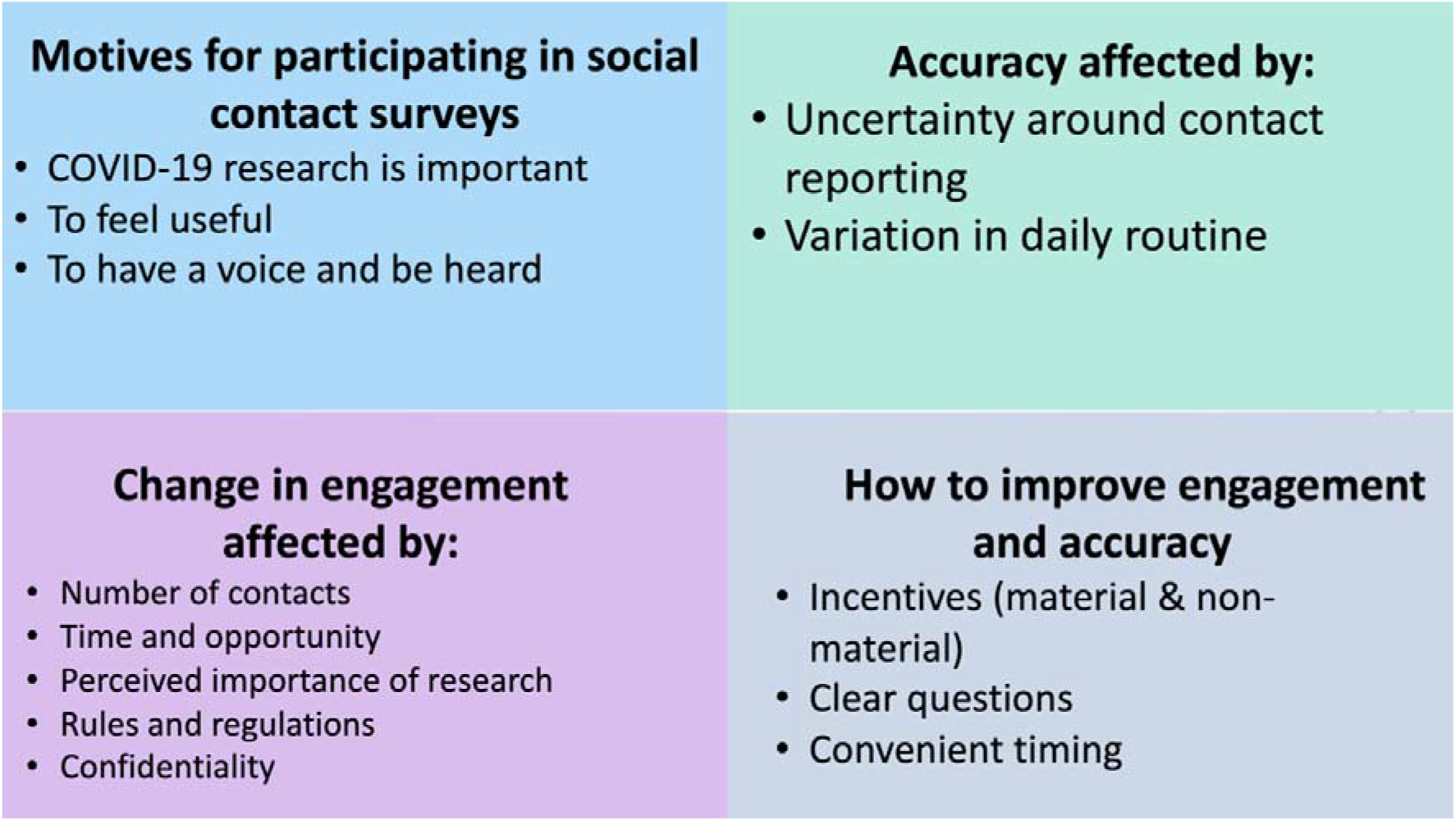
Summary of the main results of the thematic analysis of the focus group transcripts

## Discussion

### Principal findings

#### Motivations to engage with contact surveys

Students in our focus groups were motivated to participate in the CONQUEST contact survey because they viewed it as important COVID-19-related research. In accordance with existing literature, students reported taking part for a combination of altruistic reasons and for personal benefits (24,25). Their involvement provided an opportunity to contribute to the pandemic response and combat negative perceptions of students by generating an evidence-based account of their behaviours (26). Interestingly, CONQUEST participants were more likely to provide ethnicity data on recruitment than university students were on registration in 2020; this generosity with respect to personal data may reflect the perceived importance of this work. Generally, participants agreed that financial incentives and/or personalised feedback would increase participation in contact surveys.

Altruistic motivations have not been reported in relation to other student contact surveys, though none have explicitly explored this. Ninety five percent of students completing an exit survey of students following a longitudinal contact survey reported that they were motivated by financial incentives (27). One student contact survey entered participants into a lottery for a prize (28); another offered a non-material incentive in the form of a project website hosting a graphical representation of contact networks generated by the survey responses (29). The effectiveness of these incentives was not assessed.

Two contact surveys used respondent-driven or ‘chain referral’ recruitment processes, whereby early recruits (the ‘seed’ sample) were asked to nominate others to take part (27,29). Students with more nominations by existing participants (signifying that their friends and social contacts were participating) were more likely to sign up to the survey (27). Whilst this approach encourages participation from socially-connected individuals and generates important insights into contact networks, it could result in homogeneous sample that does not represent the wider student population (29).

None of the student contact surveys reviewed here reported strategies for maintaining motivation and engagement in the longer term, though attrition from longitudinal surveys was acknowledged as a problem (27,29). Our focus groups identified that small, guaranteed incentives might be more motivating than prize draws and could be awarded for repeated engagement (e.g., after completing x number of weekly surveys).

A principal reason for choosing to stop completing the survey was because participants had less time and more contacts once COVID-19 restrictions were eased. Improving the ease at which contacts can be reported in a survey is therefore an important area for future research to encourage continued participation. Alternative methods to surveys, such as the use of Bluetooth tags or personal mobile devices have also been used to collect contact data, however, some participants have reported concerns with privacy associated with these methods(30).

#### Factors influencing completion and accuracy

Survey completion was impacted by the tightening and easing of lockdown measures, which influenced how much time was spent online (i.e., greater periods online providing more opportunity to complete the survey) and the number of daily contacts (i.e., lower numbers being easier to report). These qualitative findings were supported by CONQUEST completion rates which dropped substantially when lockdown measures eased, and students were permitted to mix outside their residences. Surveys were usually completed on a weekday and in the morning, which may have corresponded with (online) teaching sessions or private study time. CONQUEST data suggest that students who regularly completed the survey were more likely to report large numbers of group contacts, compared to students who completed it as a one-off. This could be because those who completed it as a one-off were deterred from regularly completing the survey due to the time it takes to enter in contacts and may have not entered all of these (as reported by focus group members), while those who complete it regularly are more invested and engaged with the survey.

The completeness and accuracy of survey responses could, according to focus group participants, be compromised by concerns about data confidentiality and the implications of reporting lots of interactions whilst COVID-19 restrictions were in place. Participants were also concerned that responses might not truly represent student behaviours for three key reasons. Firstly, certain questions caused confusion: for example, because they did not know whether a contact should be listed as an ‘individual’ or ‘group’ contact, or because they did not have all the information requested of them. In these situations, contacts might be reported more than once or not at all. Secondly, participants reported very variable numbers of contacts from day to day; whilst some strategically chose to report on a different day each week to give the full picture, others tended to report when it fitted into their routine, or to report a quiet day with few contacts. Thirdly, participants highlighted issues with recall when reporting contacts for the previous day. These issues with completeness and accuracy of survey responses should be taken into consideration when designing future contact surveys and when using such data in epidemic modelling.

During lockdown the amount of time the UK public spent online increased as people relied on the internet for home working and schooling, shopping and communication (31) and, as our focus groups highlighted, these stay-at-home periods were conducive to completing online surveys. Other period effects, such as term times, can also affect survey completion rates (6). In addition, daily routines appear to be important: over 80% of students completing a paper contact survey in a German university did so before midday (32).

To reduce confusion when contacts are encountered in more than one context some student contact surveys have stipulated these individuals should be listed only once, for the most intimate or intense form of contact that day (28,33). One survey asked participants to enter contacts for every context/ encounter, but this instruction was unlikely to have been followed as most contacts were listed in a single context (34). Entering aggregate numbers versus individuals was not shown to affect the average number of contacts reported in a German university setting, and the authors advocate the use of a blended approach, since diaries act as a memory-aid for reporting individual contacts, whilst aggregate reporting is easier for large groups (32). Aggregate reporting is arguably necessary for university populations, as they regularly engage in group teaching and social activities. Over 40% of Belgian university survey participants joined gatherings of more than 20 adults on multiple weekdays (28).

Other student contact surveys support our observations that the number and nature (casual versus physical) of contacts varies considerably, particularly on a weekday as compared to the weekend (28,32,34,35). To combat this, surveys have deliberately been deployed on randomly assigned days (28,33,34) with oversampling on weekends in one case (34). The longitudinal survey was deployed on a Friday each week, and expired the following Tuesday (27).

With respect to recall bias, participants in German university contact surveys reported better recall for weekend contacts than weekday contacts (32). Other UK-based student contact surveys have used prospective diaries to improve contact ascertainment (33); an approach that students in Belgium found acceptable (65% reported that it required ‘little effort’) and unlikely to affect their contact behaviours (28). On comparing prospective contact reports with retrospective reports of contacts ‘yesterday’ however, this study found no significant difference in the average number of contacts reported (28). Although the German comparative study found that prospective diaries yielded slightly more contacts (on average 2 more contacts per weekday) than retrospective diaries, other metrics (duration and nature of contacts) remained consistent, and the investigators argue that the benefits of a prospective approach are outweighed by lower response rates as compared to retrospective diaries (32).

#### Strengths and limitations of this study

To our knowledge, this is the first mixed-methods evaluation of a social contact survey. The aim was to explore facilitators and barriers to completing the CONQUEST survey, highlight biases in our data that might not be immediately obvious, and identify ways to improve survey participation, data completeness and accuracy. CONQUEST was designed with the Bristol university population in mind and focuses on data collection to inform the response to the COVID-19 pandemic. We appreciate that our findings may not be generalisable to other contact surveys or student populations, though we believe that several observations may be useful in other contexts.

The utility of this study is limited by the small number of focus groups, which restricted the range of views that could be captured. The demographic profile of our focus groups was not entirely representative of CONQUEST participants, who in turn were more likely to be female and white than the wider student population. Our results may therefore not reflect the views and behaviours of the average student at the university.

Despite efforts to recruit a diverse sample of students, focus group participants and CONQUEST participants are self-selected and motivated to take part in research and our results must be interpreted with this in mind.

For the quantitative analysis of the time of day the survey was completed, we did not know the location of participants when they were completing the survey and so if students were abroad at the time of completion this will have been recorded in UK time.

#### Recommendations for improving social contact surveys

When using social contact data in epidemiological modelling, it is important to be aware of the biases we identify here, as although the data are useful, they will not be a completely accurate representation of human behaviours. Box 1 gives some recommendations from our work that could help to increase participation and improve accuracy in social contact surveys. More broadly we feel that the representativeness of UK social contact surveys could be improved; we must try harder to engage males, minority ethnic groups and international students. There may also be a role for a set of minimum standards to strengthen the validity of results from social contact surveys., which in turn will allow us to be better prepared for future epidemics and pandemics. Our findings will help to inform future contact surveys for epidemic modelling, which in turn will allow us to be better prepared for future epidemics and pandemics.

**Box 1 – Recommendations for future research and practice Encouraging ongoing participation, and a continuing sense of importance for survey completion**.

*Incentivisation*

Although students reported a range of motivations for taking part in research, incentives may be helpful for initiating and continuing completion of surveys. Small, guaranteed financial incentives may be more motivating than prize draws, although personalised feedback or communications about survey results may also be beneficial. Future research is now needed to test the efficacy of these strategies.

*Optimising contact reporting*

If possible, surveys should give the option for respondents to save the details of some ‘regular’ contacts. This would reduce frustration towards having to re-enter the details of these contacts every time and the time taken to complete the survey.

**Optimising honesty and accuracy**

*Anonymity*

Emphasizing the confidentiality of the survey is necessary to facilitate truthful reporting. Despite reporting an awareness that responses were anonymous, students were reluctant to report breaches that were considered illegal or against university procedure. Research is needed to understand the best way to ensure students feel safe to disclose possible breaches of lockdown; e.g., through providing a free-text box in which participants can justify their behaviour.

*Ambiguity*

It is important that all questions are clear, unambiguous and have a clear rationale for inclusion. Including click-through links to further information may ensure necessary information is provided to those who need it; e.g., through a “more information” button.

*Timing*

Survey deployment should be timed for maximum deployment (e.g., weekday mornings), but it is also important to consider how best to encourage completion to reflect variation in daily routines, and how to reduce attrition/ non-returns outside term time. Consider giving participants a set day to complete the survey, to reduce confusion, prevent strategic completion and capture day-to-day and week-by-week variation in contact behaviours.

**Formatting**

The assumption is that online surveys are preferred to telephone or face-to-face questioning, but this should be clarified.

## Data Availability

Data are available at the University of Bristol data repository, data.bris, at https://doi.org/10.5523/bris.29p4r41hm0oz525k33jjvevrrd, along with the code that was used for the analyses.

## Acknowledgements

We would like to thank all the participants of the focus groups for taking part, the University of Bristol student communications team for aiding with the recruitment process and our data manager, Alison Horne, for support with the CONQUEST survey data. The focus groups were funded by the Elizabeth Blackwell Institute. HC, EBP, SD, TS and RK would like to acknowledge support from the National Institute for Health Research (NIHR) Health Protection Research Unit (HPRU) in Behavioural Science and Evaluation at the University of Bristol. HC is additionally funded through an NIHR Career Development Fellowship [CDF-2018-11-ST2-015], which also funds TS. The views expressed are those of the author(s) and not necessarily those of the NIHR, the Department of Health and Social Care, or UKHSA. ATh is supported by the Wellcome Trust (217509/Z/19/Z) and UKRI through the JUNIPER consortium MR/V038613/1 and CoMMinS study MR/V028545/1. EBP, EN and AB are supported by UKRI through the JUNIPER consortium (Grant Number MR/V038613/1). EBP is further supported by MRC (Grant Number MC/PC/19067). RK is funded by the Wellcome GW4 Clinical Academic Training programme (203918). AT is supported by the Wellcome Trust (222770/Z/21/Z).

## Author contributions

EN, EBP and SD conceived the study. EN, EBP, SD and HC designed the study. EN organised the focus groups and recruitment of participants. All authors were involved in the running of the focus groups. TS and SD did the qualitative analysis. EN, AT and AB did the quantitative analysis. EN, EBP, SD, TS, AB, AT and RK wrote the first draft of the paper. All authors were involved in interpretation of the results, revising the paper and approving the final version to be submitted.

## Data statement

Data are available at the University of Bristol data repository, data.bris, at https://doi.org/10.5523/bris.29p4r41hm0oz525k33jjvevrrd.

## References

1. Hens N, Ayele GM, Goeyvaerts N, Aerts M, Mossong J, Edmunds JW, et al. Estimating the impact of school closure on social mixing behaviour and the transmission of close contact infections in eight European countries. BMC Infectious Diseases. 2009 Nov;9(1):187.

2. Edmunds WJ, O’Callaghan CJ, Nokes DJ. Who mixes with whom? A method to determine the contact patterns of adults that may lead to the spread of airborne infections. Proceedings of the Royal Society B: Biological Sciences. 1997;264(1384):949–57.

3. Hoang T, Coletti P, Melegaro A, Wallinga J, Grijalva CG, Edmunds JW, et al. A Systematic Review of Social Contact Surveys to Inform Transmission Models of Close-contact Infections. Epidemiology. 2019 Sep;30(5):723–36.

4. Mossong J, Hens N, Jit M, Beutels P, Auranen K, Mikolajczyk R, et al. Social contacts and mixing patterns relevant to the spread of infectious diseases. PLoS medicine. 2008;5(3):e74.

5. Danon L, Read JM, House TA, Vernon MC, Keeling MJ. Social encounter networks: characterizing Great Britain. Proceedings Biological sciences / The Royal Society. 2013 Aug;280(1765):20131037.

6. Hoang T, Coletti P, Melegaro A, Wallinga J, Grijalva CG, Edmunds JW, et al. A Systematic Review of Social Contact Surveys to Inform Transmission Models of Close-contact Infections. Epidemiology. 2019 Sep;30(5):723–36.

7. Edmunds WJ, O’Callaghan CJ, Nokes DJ. Who mixes with whom? A method to determine the contact patterns of adults that may lead to the spread of airborne infections. Proceedings of the Royal Society B: Biological Sciences. 1997;264(1384):949–57.

8. Beutels P, Shkedy Z, Aerts M, van Damme P. Social mixing patterns for transmission models of close contact infections: exploring self-evaluation and diary-based data collection through a web-based interface. Epidemiology & Infection. 2006 Dec;134(6):1158–66.

9. Edmunds WJ, Kafatos G, Wallinga J, Mossong JR. Mixing patterns and the spread of close-contact infectious diseases. Emerging Themes in Epidemiology [Internet]. 2006 Aug 14 [cited 2022 Jan 22];3(1):1–8. Available from: https://ete-online.biomedcentral.com/articles/10.1186/1742-7622-3-10

10. Danon L, Read JM, House TA, Vernon MC, Keeling MJ. Social encounter networks: characterizing Great Britain. Proceedings Biological sciences / The Royal Society. 2013 Aug;280(1765):20131037.

11. Baltrusaitis K, Santillana M, Crawley AW, Chunara R, Smolinski M, Brownstein JS. Determinants of Participants’ Follow-Up and Characterization of Representativeness in Flu Near You, A Participatory Disease Surveillance System. JMIR Public Health Surveill 2017;3(2):e18 https://publichealth.jmir.org/2017/2/e18. 2017 Apr;3(2):e7304.

12. Bajardi P, Vespignani A, Funk S, Eames KTD, Edmunds WJ, Turbelin C, et al. Determinants of Follow-Up Participation in the Internet-Based European Influenza Surveillance Platform Influenzanet. J Med Internet Res 2014;16(3):e78 https://www.jmir.org/2014/3/e78. 2014 Mar;16(3):e3010.

13. Baltrusaitis K, Santillana M, Crawley AW, Chunara R, Smolinski M, Brownstein JS. Determinants of Participants’ Follow-Up and Characterization of Representativeness in Flu Near You, A Participatory Disease Surveillance System. JMIR Public Health Surveill 2017;3(2):e18 https://publichealth.jmir.org/2017/2/e18. 2017 Apr;3(2):e7304.

14. Bajardi P, Paolotti D, Vespignani A, Eames K, Funk S, Edmunds WJ, et al. Association between Recruitment Methods and Attrition in Internet-Based Studies. PLOS ONE. 2014 Dec;9(12):e114925.

15. Mastrandrea R, Fournet J, Barrat A. Contact Patterns in a High School: A Comparison between Data Collected Using Wearable Sensors, Contact Diaries and Friendship Surveys. PLOS ONE. 2015 Sep;10(9):e0136497.

16. Beutels P, Shkedy Z, Aerts M, Van Damme P. Social mixing patterns for transmission models of close contact infections: exploring self-evaluation and diary-based data collection through a web-based interface. Epidemiology & Infection. 2006 Dec;134(6):1158–66.

17. Nixon E, Trickey A, Christensen H, Finn A, Thomas A, Relton C, et al. Contacts and behaviours of university students during the COVID-19 pandemic at the start of the 2020/2021 academic year. Scientific Reports 2021 11:1. 2021 Jun 3;11(1):1–13.

18. Trickey A, Nixon E, Christensen H, Finn A, Thomas A, Relton C, et al. University students and staff able to maintain low daily contact numbers during various COVID-19 guideline periods. Epidemiology & Infection. 2021;149.

19. Creswell JW, Creswell JD. Mixed Methods Procedures. In: Cresswell JW, Cresswell JD, editors. Research Design: Qualitative, Quantitative, and Mixed M ethods Approaches. 5th ed. SAGE Publications; 2018. p. 418.

20. Morgan DL. Focus Groups. https://doi.org/101146/annurev.soc221129. 2003 Nov 28;22:129–52.

21. Barbour R, Kitzinger J. Developing Focus Group Research. In: Developing Focus Group Research. SAGE Publications Ltd; 1999. p. 1–20.

22. Braun V, Clarke V. Reflecting on reflexive thematic analysis. https://doi.org/101080/2159676X20191628806. 2019 Aug 8;11(4):589–97.

23. University of Bristol UK. Student numbers and demographics | Education Administration Office | University of Bristol [Internet]. [cited 2022 Jan 21]. Available from: https://www.bristol.ac.uk/ssio/statistics/

24. Manton KJ, Gauld CS, White KM, Griffin PM, Elliott SL. Qualitative study investigating the underlying motivations of healthy participants in phase I clinical trials. [cited 2022 Jan 27]; Available from: http://bmjopen.bmj.com/

25. Locock L, Smith L. Personal benefit, or benefiting others? Deciding whether to take part in clinical trials. Clinical Trials [Internet]. 2011 Feb 16 [cited 2022 Jan 27];8(1):85–93. Available from: https://journals.sagepub.com/doi/10.1177/1740774510392257?url_ver=Z39.88-2003

26. Clark T. On ‘being researched’: why do people engage with qualitative research? Qualitative Research. 2010;10(4).

27. Aiello AE, Simanek AM, Eisenberg MC, Walsh AR, Davis B, Volz E, et al. Design and methods of a social network isolation study for reducing respiratory infection transmission: The eX-FLU cluster randomized trial. Epidemics. 2016 Jun 1;15:38–55.

28. Beutels P, Shkedy Z, Aerts M, Van Damme P. Social mixing patterns for transmission models of close contact infections: exploring self-evaluation and diary-based data collection through a web-based interface. Epidemiology & Infection. 2006 Dec;134(6):1158–66.

29. Stein ML, Van Steenbergen JE, Buskens V, Van Der Heijden PGM, Chanyasanha C, Tipayamongkholgul M, et al. Comparison of Contact Patterns Relevant for Transmission of Respiratory Pathogens in Thailand and the Netherlands Using Respondent-Driven Sampling. PLOS ONE. 2014 Nov 25;9(11):e113711.

30. Shelby T, Caruthers T, Kanner OY, Schneider R, Lipnickas D, Grau LE, et al. (No Title). Available from: https://formative.jmir.org/2021/10/e31086

31. Ofcom. Online Nation 2021 report [Internet]. 2021 [cited 2022 Jan 21]. Available from: https://www.ofcom.org.uk/data/assets/pdf_file/0013/220414/online-nation-2021-report.pdf

32. Mikolajczyk RT, Kretzschmar M. Collecting social contact data in the context of disease transmission: Prospective and retrospective study designs. Social Networks. 2008 May 1;30(2):127–35.

33. Edmunds WJ, Kafatos G, Wallinga J, Mossong JR. Mixing patterns and the spread of close-contact infectious diseases. Emerging Themes in Epidemiology. 2006 Aug 14;3(1):1–8.

34. Edmunds WJ, O’Callaghan CJ, Nokes DJ. Who mixes with whom? A method to determine the contact patterns of adults that may lead to the spread of airborne infections. Proceedings of the Royal Society B: Biological Sciences. 1997;264(1384):949–57.

35. Read JM, Eames KTD, Edmunds WJ. Dynamic social networks and the implications for the spread of infectious disease. Journal of the Royal Society Interface. 2008 Sep 6;5(26):1001.

