## Supplementary materials for "A mixed methods analysis of participation in social contact surveys"

### Supplementary material for “A mixed methods analysis of participation in social contact surveys

**Supplement 1: Topic guide for CONQUEST focus groups**

**Students who have completed CONQUEST** **(Groups 1 and 2)**

***Enrolment and motivation***

- How did you find out about the survey?
- What made you decide to take part in the survey?
- What made you stop taking part (if you haven’t taken part in a while)?

***Completing the survey***

- [If you do] Why do you continue to complete the survey?
- [If you don’t] Why do you not continue to complete the survey?
- Are there particular times of day or week that you are more likely to complete the survey?
- Are there any factors that might impact the likelihood of you completing the survey (prompts: having a lot of contacts on the previous day, symptoms, a recent test)?
- Do you usually complete the survey on your phone, tablet or computer? (Any issues with these?)
- Have you ever only completed part of the survey (dropped out before finishing)? If so, why?

***The survey***

- Which questions / parts of the survey work well?
- Which parts of the survey are the most difficult to complete?
- What could we do to make it more appealing?
- What could we do to stop people dropping out of the study (not completing it after the first attempt)?
- What could we do to make it easier for other people to complete the survey?

**Go through the contacts question in detail (Supplement 2)**

- Is there anything you don’t think is clear about this question?
- How do they interpret the question (does everyone interpret it in the same way)?
- Is there anything that would stop you responding honestly to this question?

**Students who have not completed CONQUEST** **(Groups 3 and 4)** 

***Enrolment and motivation***

- Have you heard of the survey?
- [If yes] Why did you decide not to take part?
- What would motivate you to take part?
- Do you take part in any other research?
- What could we do to encourage more students to take part in the survey?

***Completing the survey***

- Are there particular times of day or week that you are more likely to complete the survey?
- How often do you think you would complete the survey?
- Are there any factors that might impact the likelihood of you completing the survey? (prompts: having a lot of contacts on the previous day, symptoms, a recent test)

 

**Go through the contacts question in detail (Supplement 2)**

- Is there anything you don’t think is clear about this question?
- How do they interpret the question (does everyone interpret it in the same way)?
- Is there anything that would stop you responding honestly to this question?

**Supplement 2: Contacts question**

**Section 6 – Your contacts yesterday**

This section is asking about the contacts that you had with other people **yesterday including those that you live with.**

There are three parts to this section of the survey about three different types of contacts you might have had. Read the following instructions carefully so that you know where to enter each contact.

(1) Individual contacts- those who you spoke in person to one-on-one, including those in your household and support bubble
(2) If you spoke in person to many people one-on-one in the same setting (but they did not have the opportunity to speak to each other), for example, as part of working in a customer service role in a shop
(3) Large groups of individuals in the same setting (e.g. sports teams, tutorials, lectures, religious services, large gatherings with friends and family)

PLEASE RECORD EACH PERSON ONLY ONCE, on a single row, or as a member of just one group, even if you meet the same person several times during the day and/or as part of a group. If you spoke to an individual in multiple locations, choose the location where you spent the most time with that person.

This survey is anonymous – **it is important for you to record all of your contacts and how they happened, whether they fit with the current social/physical distancing advice or not**.

***Individual contacts yesterday***

Thinking about yesterday, who did you speak to **(in person) yesterday**? Only include individuals you spoke to one-on-one as you will be able to enter those you met in a group in a later section. Do include those that are a part of your household.

It can help to think about your day from when you got up, at breakfast, in the morning, at lunch, in the afternoon, at teatime, and in the evening.

Please include everyone you have been close to. This includes:

- People you had a face to face conversation without a complete barrier between you. Please include people you were within 3 meters (10 feet) of and spoke to. Do not include people you talked to through a solid barrier, for example, a closed window,
- People you had physical contact with for example, handshake, hug, kiss.
- Examples of these contacts could be your partner, a parcel delivery person, a friend or a shop worker
- Only include individuals you spoke to one-on-one as you will be able to enter those you met in a group in a later section.
- Do include those that are a part of your household
- If you spoke to many people one-on-one in the same setting (but they did not have an opportunity to speak to each other), for example, as part of a customer service role, you are not required to enter these particular contacts separately but will be prompted to include these in a later question.
- If you spoke to an individual in multiple locations, choose the location where you spent the most time with that person.

I did not speak to anyone in person yesterday on a one-on-one basis [*can skip to next section if selected]*

*If you have done this survey before, the answers from the last survey you completed are shown below. Please check these carefully and change your responses if they are different. If there has been no change then please select at the bottom of the page “No changes”.*

Person 1:

Name (or description):

- Age: 0-4/ 5-17/18-24/25-44/ 45-64/ 65-80/81+

Is this person part of your household? Yes/No

Does this person work or study at the University of Bristol? Yes/No/Don’t know

What is the main faculty this person is associated with? Arts/Engineering/Health Sciences/ Life Sciences/Science/Social Sciences/Law/Other/Don’t know

Which school are they in?

Did you also touch this person? Yes/No

Where were you when you spoke to this person? Home/Another home /University/Work or volunteering not at university/School or nursery/Shopping/Medical or health centre/Transport/Place of worship/Social (pub/nightclub/café/restaurant)/Exercise/Park/Other location

Was this inside, outside or both?

How long did you talk to this person for? Less than 10 minutes / Between 10 minutes and an hour / between 1 and 4 hours / 4+ hours

How often would you expect to meet this person (or one person providing this service if applicable) under current physical distancing measures? 4 or more days a week/2-3 days a week/Once a week/Less often than once a week/Met for the first time this day

Repeat for all people you spoke to yesterday, until you click “I was in contact with no more people”

I was in contact with no more people []

**Additional conversational or physical contacts (e.g. as part of a customer service role)**

If you had any additional individual contacts where you had a conversation with a person or had physical contact, please enter the number of contacts below.  For example, if you work in a shop and had conversations with 24 customers in a day, enter 24.  If you do not know the exact number, please enter your best estimate.

| 14 | Number of additional individual conversational/ physical contacts   [  ][  ] [] [] |
| --- | --- |

***Group contacts yesterday***

In addition to the people you listed in question 4, did you meet with any large groups **yesterday** (for example, sports teams, tutorials, lectures, religious services, large gatherings with friends and family)?

- I didn’t meet with any groups yesterday [*if selected then the next question does not appear]*

Group 1:

Name (or description):

How many people were in the group in each age category?

- Age 0-4:
- Age 5-17:
- Age 18-24:
- Age 25-44:
- Age 45-64:
- Age 65-80:
- Age 81+:

Do the majority of this group work or study at the University of Bristol? Yes/No

What is the main faculty this group is associated with? Arts/Engineering/Health Sciences/ Life Sciences/Science/Social Sciences/Law/Other/Don’t know

What is the main school this group is associated with?

Where were you when you spoke to these people? Home/Another home /University/Work or volunteering not at university /School and nursery/Shopping/Medical or health centre /Transport/Place of worship/Social (pub/nightclub/café/restaurant)/exercise/park/other location

Was this inside or outside?

How long did you talk to this group for? Less than 10 minutes / Between 10 minutes and an hour /Between 1 and 4 hours / 4+ hours

Did the members of the group also talk to each other?

How long did you talk to this group for? Less than 10 minutes / Between 10 minutes and an hour / between 1 and 4 hours / 4+ hours

How often would you expect to meet this group under current physical distancing measures? 4 or more days a week/2-3days a week/Once a week/Less often than once a week/Met for the first time this day

Repeat for all groups you spoke to yesterday, until you click “I was in contact with no more groups”

I was in contact with no more groups []
